# The Burden and Transmission Dynamics of Toxoplasmosis in Relation to Congenital Diseases among Pregnant Women in Ghana

**DOI:** 10.1101/2024.12.04.24318473

**Authors:** Ebenezer Assoah, Denis Dekugmen Yar, Papa Kofi Amissah-Reynolds, Gadafi Iddrisu Balali, Rockson Addy, Joshua Kpieonuma Zineyele

**Affiliations:** Department of Biological Sciences Education, Akenten Appiah-Menka University of Skills Training and Entrepreneurial Development (AAMUSTED), Asante Mampong Campus, Ghana; Department of Public Health Education, Akenten Appiah-Menka University of Skills Training and Entrepreneurial Development (AAMUSTED), Asante Mampong Campus, Ghana; Department of Theoretical and Applied Biology, Kwame Nkrumah University of Science and Technology, Kumasi, Ghana; Department of Science Education, Seventh–Day Adventist College of Education, Agona-Ashanti, Ghana; Department of Science and Information Communication Technology, Effiduase Senior High Technical School, Ashanti Region, Ghana

**Keywords:** *Toxoplasma gondii*, Transmission Dynamics, Seroprevalence, Congenital Diseases, Asante-Mampong

## Abstract

This study assessed the Burden and Transmission Dynamics of Toxoplasmosis in Relation to Congenital Diseases among Pregnant Women in the Asante Mampong Municipality of Ghana. A cross-sectional design was used to recruit 201 pregnant women from six health facilities conveniently. Participants’ socio-demographics, clinical and environmental data were collected using a structured questionnaire. Using 2ml of blood, *T. gondii* seroprevalence was determined by the TOXO IgG/IgM Rapid Test Cassette. Data was analyzed using descriptive and logistic regression analysis with SPSS version 27 to determine the prevalence and associations of *T. gondii* infection with other variables, respectively. The seroprevalence of *T. gondii* was 49.75%, of which 40.30%, 2.49%, and 6.97% tested positive for IgG, IgM, and IgG/IgM, respectively. Co-infection of toxoplasmosis with viral hepatitis B, HIV, and syphilis rates were 15%, 1%, and 4%, respectively and were not risk factors for *T. gondii* transmission. Educational level and residential status were associated with toxoplasmosis [p<0.05]. Participants with higher education had a reduced risk of *T. gondii* infections compared to a lower level of education [AOR= 0.39 (0.13, 0.99) p=0.049]. Similarly, those residing in peri-urban and urban areas had a reduced risk of infection with *T. gondii* [AOR= 0.13 (0.02, 0.7) p=0.02] and [AOR= 0.10 (0.02, 0.78) p=0.03], respectively. Backyard animals with extensive and semi-intensive systems, without veterinary care, and contact with animal droppings and water sources were significant risk factors for *T. gondii* infection [p<0.05]. Miscarriage was associated with *T. gondii* infection [p<0.05]. The burden of *T. gondii* infection was high among the study population, with the risk of mother-child transmission. Level of education, residence, backyard animal farms, hygiene practices, water sources and quality were risk factors for *T. gondii* infection. Toxoplasmosis is a risk factor for miscarriage, and therefore, integrating it into ANC routine screening could improve pregnancy outcomes.

**Author Summary:** Toxoplasmosis, caused by the parasite *Toxoplasma gondii*, is a significant health problem, particularly for pregnant women due to its potential to cause severe congenital diseases. This study examined the prevalence and risk factors of toxoplasmosis among pregnant women in the Asante Mampong Municipality of Ghana. Nearly half of the study participants tested positive for *T. gondii*, showing a substantial public health burden. The analysis identified several contributing factors, including lower educational levels, rural residency, backyard farming practices, and poor water sources, all of which increased susceptibility to infection. Miscarriage was also associated with toxoplasmosis. The study demonstrates the need for integrating routine toxoplasmosis screening into antenatal care to mitigate adverse pregnancy outcomes and reduce transmission risks. This research provides vital data that could inform public health policies to improve maternal and neonatal health in Ghana.

## Introduction

Globally, parasitic zoonoses are on the rise, affecting more than 4 billion people, with over 200,000 deaths recorded annually [1, 2]. In low-income countries, neglected parasitic diseases such as ascariasis, amebiasis, giardiasis, toxoplasmosis, etc., continue to cause significant public health challenges, with enormous morbidity and mortality [3]. *Toxoplasma gondii* is known to infect vertebrate animals and humans [4, 5], with 25.7% and 33.8% reported among the general human population and pregnant women, respectively [6, 7]. The most significant burden is in developing countries, linked to poor sanitation and a lack of quality health services [7–10]. Children and pregnant women are generally more vulnerable to *T. gondii* infection due to their weaker immunity [11]. Pregnant women infected with *T. gondii* in the first trimester can invade the placenta, causing inflammation and damage to arteries and veins that may block the flow of blood and nutrient exchange between the mother and fetus, resulting in spontaneous abortion [12]. Meanwhile, *T. gondii* infection in the second and third trimesters may cause congenital toxoplasmosis, leading to mental retardation, auditory defects, and chorioretinitis [13, 14].

In Ghana, policies for screening expectant mothers exist for a range of congenital diseases, such as viral hepatitis, HIV/AIDS, syphilis and malaria, to improve pregnancy outcomes and neonatal health [15, 16]. The prevalence of toxoplasmosis among pregnant women in Ghana is among the highest in the world (81-90%), with possible dire consequences on pregnancy outcomes [7, 17]. Nevertheless, there is no policy for *T. gondii* screening during pregnancy in the country. However, in Europe (Austria, France, and Slovenia), mandatory national policies for prenatal serological screening and treatment for *Toxoplasma gondii* infection have significantly reduced congenital toxoplasmosis [18]. Early detection and administration of anti-*Toxoplasma* prophylaxis during pregnancy prevent congenital transmission and reduce sequelae in neonates [18].

Over the past two decades, significant progress has been made in maternal health in Ghana. Notwithstanding, maternal and neonatal morbidity and mortality are still on the increase, attributable to failure to screen and treat these diseases, including toxoplasmosis [19].

Several studies have assessed the prevalence of *Toxoplasma gondii* infection among expectant mothers, with limited focus on its comorbidity with congenital diseases and its transmission dynamics in Ghana [17, 20]. Meanwhile, data on miscarriages among pregnant women over the last decade in the Mampong Municipality is on the rise [21]. However, the proxy determinants of the high miscarriage rates are uncertain and could be linked to toxoplasmosis and other congenital diseases [22].

There is limited literature on the causal relationship between transmission dynamics and co-infection of *Toxoplasma gondii* with HIV, viral hepatitis B, and syphilis in pregnancy in the Municipality. Hence, this study assessed the burden and co-infection of *Toxoplasma gondii* with HIV, viral hepatitis B, and syphilis and its transmission dynamics among pregnant women in the Mampong Municipality of Ghana.

### Toxoplasmosis Burden, Transmission Dynamics and its Impact

Figure 1 shows the key factors contributing to the burden of Toxoplasmosis and the pathway towards its reduction. Risk factors, demographic features, and co-infections collectively increase the burden of Toxoplasmosis, which subsequently leads to various health effects. Interventions such as creating awareness, improving healthcare access, and mass screening aim to reduce this burden, with the ultimate goal of achieving zero Toxoplasmosis [23–25]

**Figure 1:**
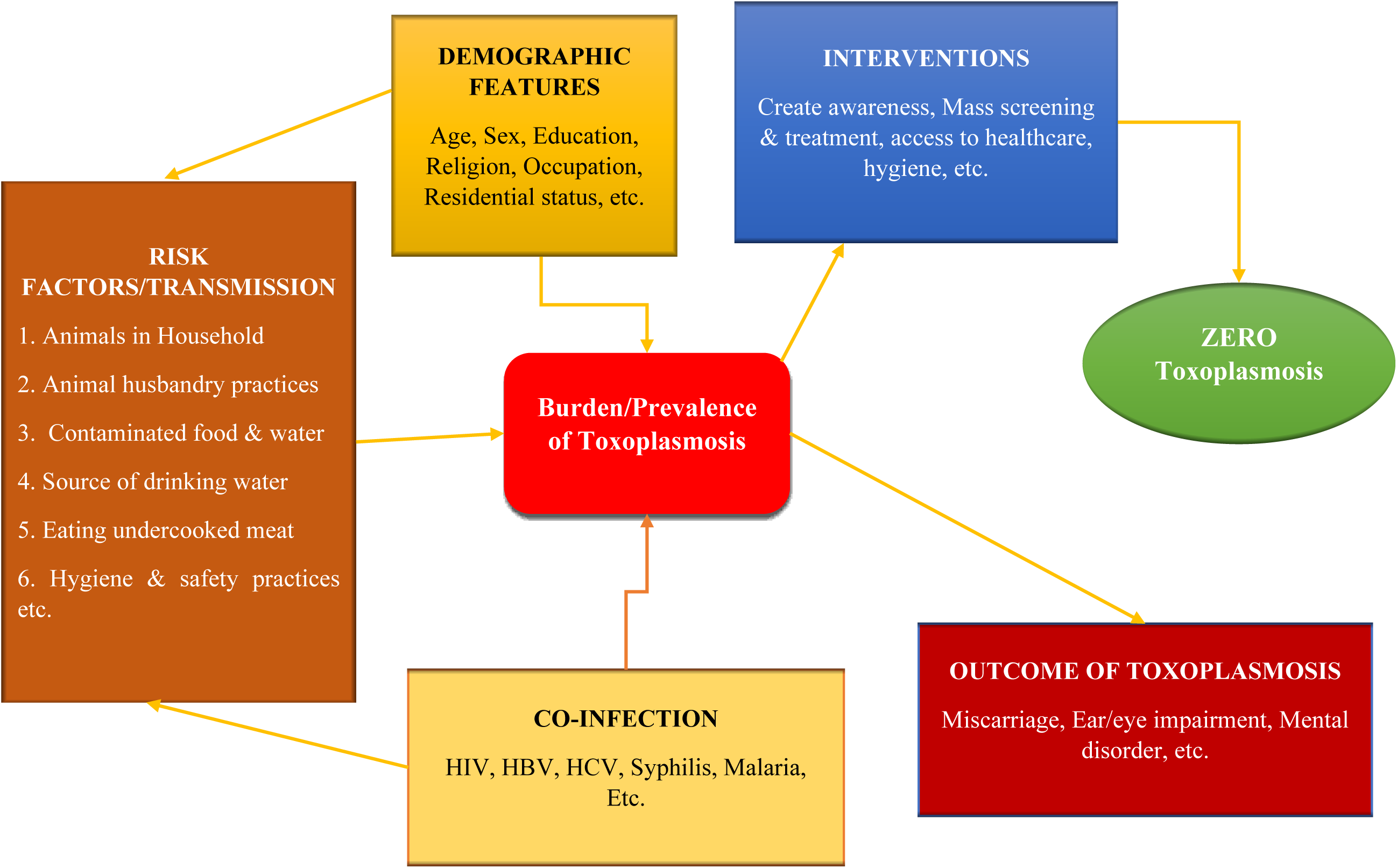
Factors contributing to the burden of Toxoplasmosis and the pathway towards its reduction

### DEMOGRAPHIC FEATURES

Age, Sex, Education, Religion, Occupation, Residential status, etc.

## 2. Materials and Methods

### Study Design

This was a facility-based cross-sectional study that assessed the burden and concurrence of *Toxoplasma gondii* infection with HBV, HIV, and syphilis among pregnant women attending antenatal care (ANC) services in health facilities. The risks of its transmission were also assessed in the Mampong Municipality from August 2023 to May 2024.

### Study Area

Figure 2 is the map of the Asante Mampong Municipal and the study sites. The study was conducted in six health facilities in five communities, with Mampong as its capital town, Dadease, Krobo, Asaam, and Kofiase. The municipality has a total population of 116,632, with females slightly more than males [26]. The municipality has an average temperature of 28°C with a relative humidity of 63% [26]. The main economic activity in the municipality is agricultural activities, including animal farming, engaging around 67.30% of the workforce [26, 27]. Most households in the Municipality own domesticated animals, such as sheep, goats, poultry, cattle, cats, and dogs.

**Figure 2:**
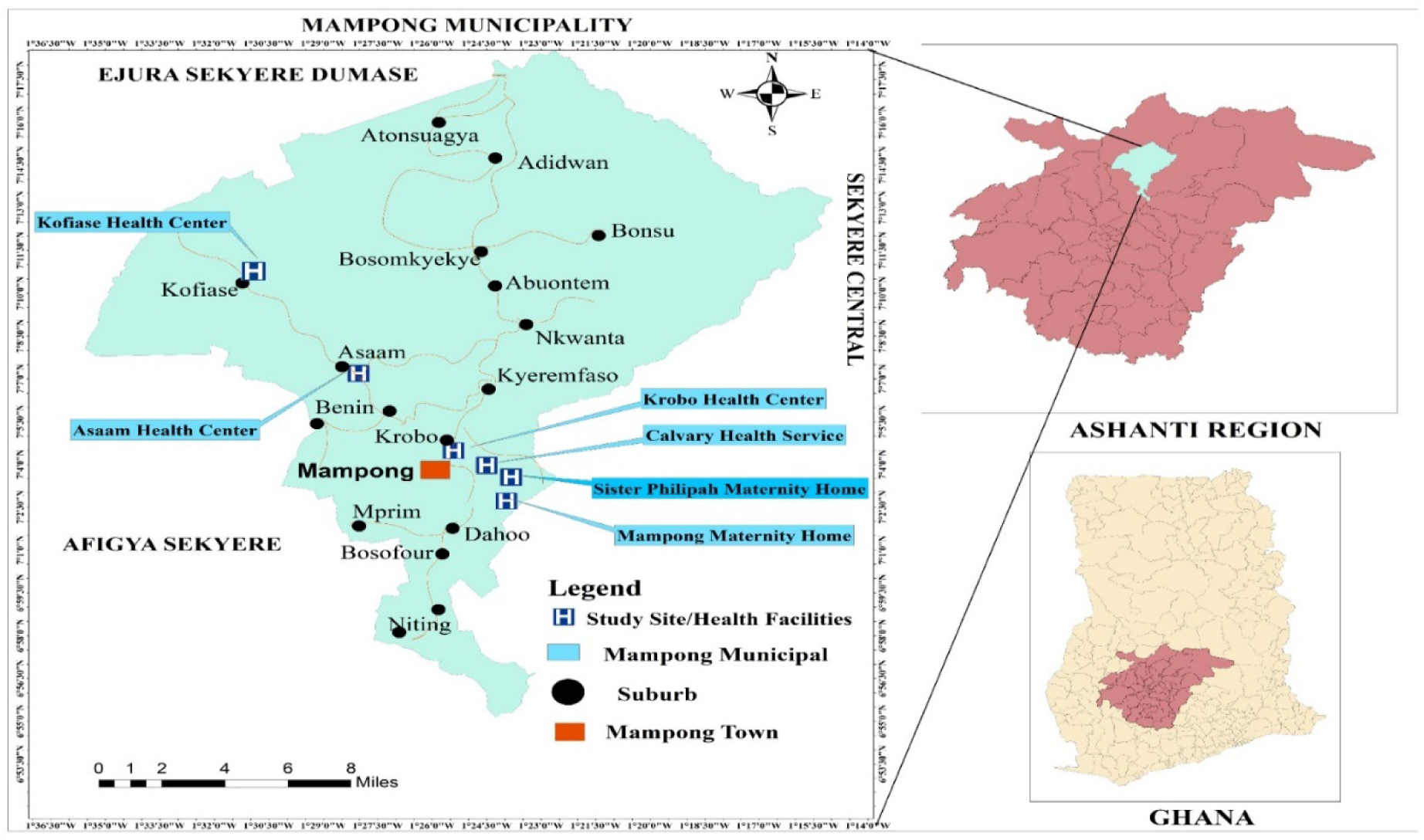
A Map of Mampong Municipality showing the Health Facilities or Study Sites

### Study Site

Data for the study were collected from the various health facilities with maternal and laboratory departments in the municipality. Mampong Government Hospital-Maternity Home, Calvary Health Center, Sister Phillipah Maternity Home and Clinic, Krobo-Dadease Health Center, Asaam Health Center, and Kofiase Health Center were used for the study, which has most pregnant women attending ANC. These facilities were purposefully chosen based on their distinct characteristics that would ensure balanced urban and rural data to reduce bias. The facilities were categorized into government and private hospitals, health centres, and clinics. This approach consequently yielded a substantial dataset suitable for making inferences.

### Study Population and Sample Size

Pregnant women of reproductive age (15 to 49 years) visiting the six selected health facilities for prenatal care and related services in the Mampong municipality were included in the study. Participants who had undergone laboratory and ultrasonography tests confirming their pregnancy and had lived in the study area for at least six months were recruited for the study. A sample size of 201 was estimated based on the toxoplasmosis prevalence rate of 92%, using Cochran’s formula as previously described [28].

### Data Collection Tools and Techniques

A structured questionnaire consisted of four sections, including socio-demographic characteristics such as age, marital status, religion, education, employment status, occupation, and residential status; obstetric variables such as gestational stage and gravida; laboratory test results such as HBV, HIV, and syphilis; and environmental variables such as water sources and the presence and confinement levels of backyard farm animals, and pets. The study participants attending antenatal care were conveniently sampled at all the selected health facilities.

### Data Collection Procedure

After obtaining informed consent, face-to-face interviews and observational methods were employed to obtain responses from the participants using a structured questionnaire. Furthermore, routine screening tests for pregnant women attending antenatal clinics were extracted from their ANC record books.

### Blood Sample Collection and Laboratory Methods

A 2ml of each participant’s blood was collected using venipuncture and into EDTA tubes. This was centrifuged at 1780 x g for 10 minutes at 4^0^C to separate plasma for subsequent serological analysis. The plasma was promptly assayed for anti-*T. gondii* IgG and IgM antibodies using the immunochromatography Test (ICT) technique. The specific ICT used was the TOXO IgG/IgM One-Step Rapid Test Cassette (WB/S/P) produced by Evancare Medical (Nantong) Co., Ltd. (China). This assay employs a lateral flow chromatographic immunoassay for the simultaneous detection and differentiation of IgG and IgM anti-*T. gondii* in human serum, plasma, or whole blood. Testing procedures were strictly followed according to the manufacturer’s instructions [29].

### Statistical Analysis

Microsoft Excel (version 2016) software was used for the data entry and exported into IBM SPSS version 27.0 for statistical analysis. Descriptive statistics were employed to determine the frequency and percentages of *T. gondii*, HBV, HIV, syphilis, and other variables. Logistics regression was used to determine the concurrence of toxoplasmosis with congenital diseases and its association with the socio-demographics and environmental variables at 0.05 and a 95% confidence interval.

### Ethical Review and Clearance

Ethical clearance was obtained from the Committee on Human Research, Publications, and Ethics (CHRPE) at the Kwame Nkrumah University of Science and Technology (CHRPE/AP/717/23). Written permissions were obtained from the Ashanti Regional and Asante Mampong Municipal Health Directorates and the study facilities. A signed informed consent form was obtained from each study participant before sample collection.

## 3. Results

### Seroprevalence of *T. gondii*

In Figure 3, th**e** prevalence of *T. gondii* infection among the participants was 49.75%, of which 40.30%, 2.49%, and 6.97% tested positive for IgG, IgM, and IgG/IgM, respectively.

**Figure 3:**
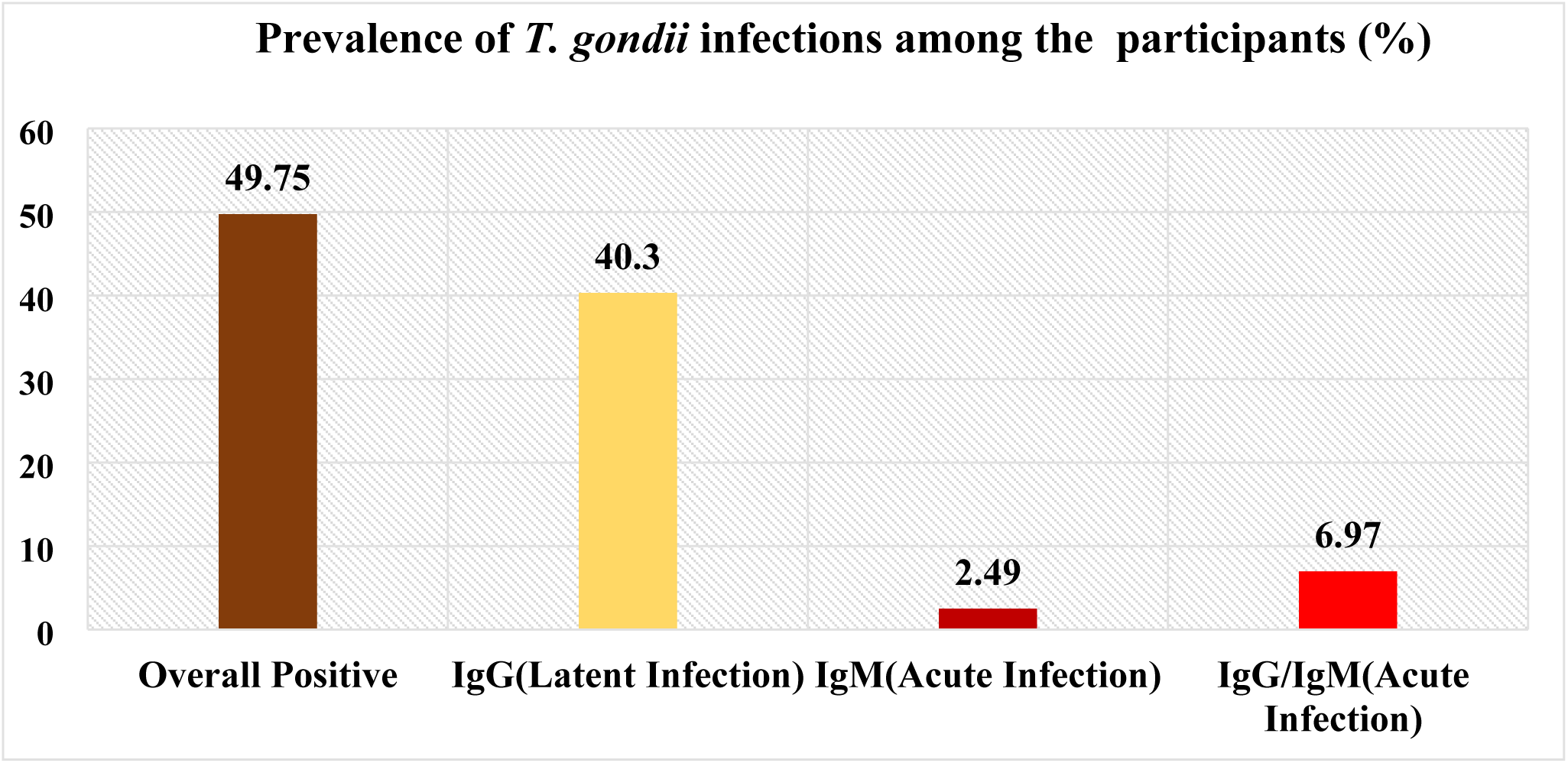
Seroprevalence of T. gondii infection in the study participants

### Prevalence of Congenital Infections

In **Table 1**, the prevalence of some congenital infections screened at the ANC was 14.61%, 0.61% and 3.07% for HBV, HIV and syphilis, respectively. There were variations in the sample sizes occasioned by the non-availability of routine screening tests at the ANC clinics for these routine ANC tests.

**Table 1:**
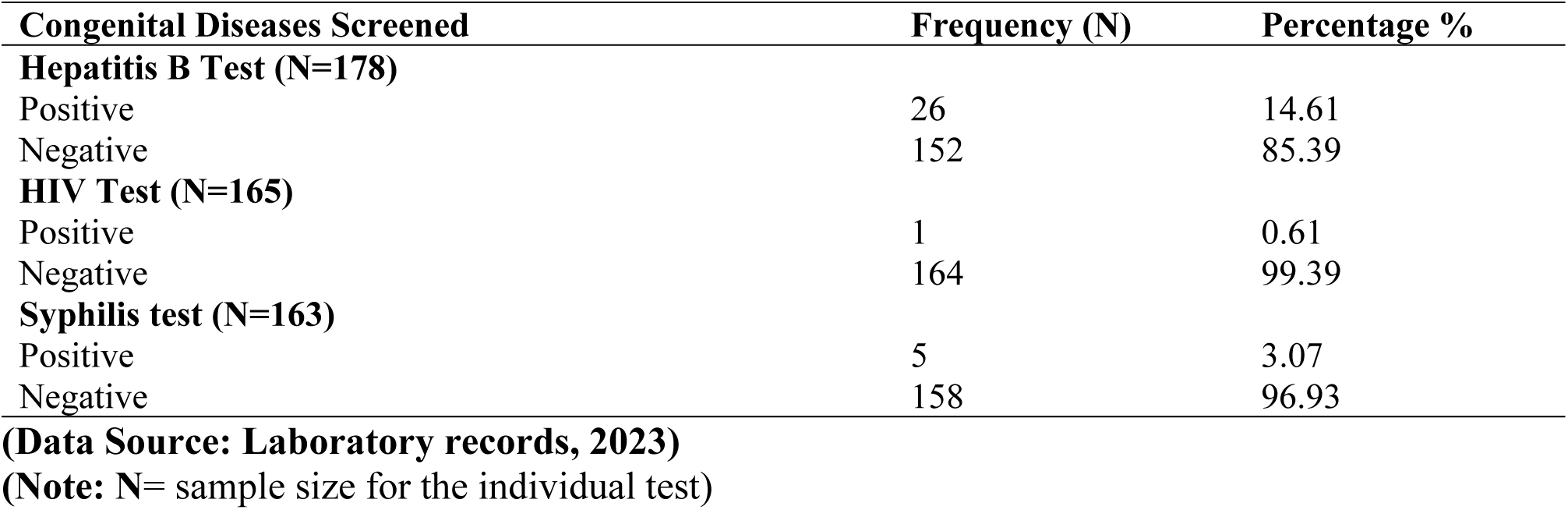
Prevalence of some Congenital Diseases Screened at ANC.

### Toxoplasmosis Concurrence with Congenital Infections Screened among Participants

**Table 2** shows the prevalence of co-infection of toxoplasmosis with HBV, HIV, and syphilis were 15%, 1%, and 4%, respectively. *T. gondii* infection was not associated with the congenital diseases screened at ANC (**P-value > 0.05).**

**Table 2:**
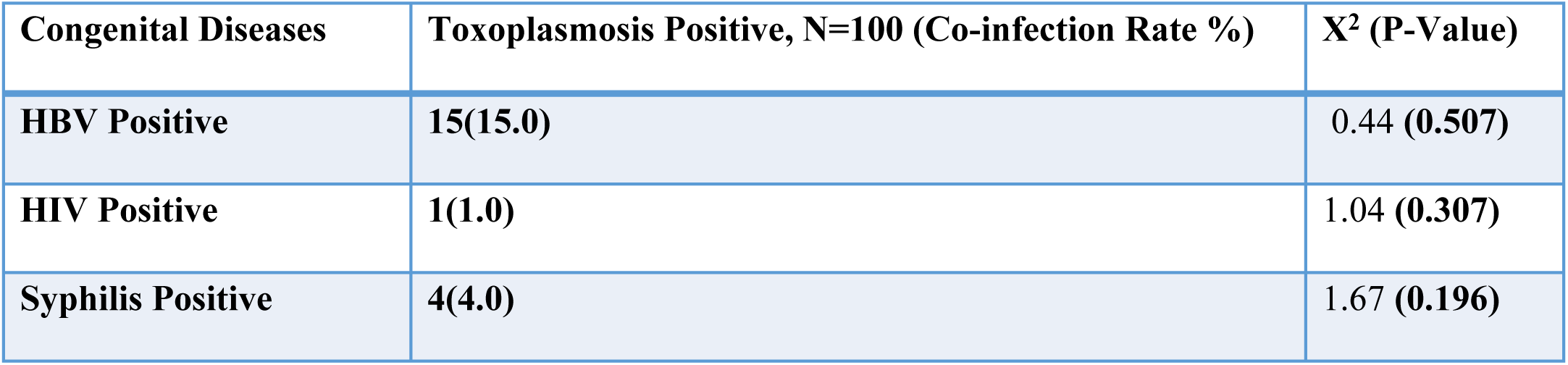
Toxoplasmosis Concurrence with Congenital Infections Screened at ANC (Data Source: Laboratory records, 2023) Note: Co-Infection (Number of positives for ANC diseases/Total number of positives for *T. gondii*) × 100%.

### Association of *T. gondii* infection with sociodemographic characteristics of participants

In **Table 3**, the level of education and residential area were significantly associated with *T. gondii* infection (χ^2^=6.56, p=0.04) and (χ^2^=10.97, p=0.004), respectively. Participants with tertiary education were less likely to be infected with *T. gondii* infections than those with lower levels of education [AOR=0.39 (0.13 – 0.99) p=0.049]. Expectant mothers who reside in the peri-urban and urban areas were less likely to be infected with *T. gondii* as compared with rural residents [AOR=0.13 (0.02 –0.70) p=0.02; AOR=0.10 (0.02 – 0.78) p=0.03]. Age, religion, gestational period, and gravida were not associated with toxoplasmosis (**P-value > 0.05).**

**Table 3:** Association of toxoplasmosis with Participants’ Socio-demographic characteristics.

| Demographics | *T. gondii* Positive (%) | $\chi^2$ (P- v) | COR (95%CI) p-v | AOR (95%CI) p-v |
| --- | --- | --- | --- | --- |
| **Age range** |  |  |  |  |
| 15 to 19 | 18 (58.1) | 1.71 (0.43) | Ref | Ref |
| 20 to 30 | 49 (51.0) |  | 0.68 (0.25, 1.34) 0.21 | 0.68 (0.26, 1.74) 0.42 |
| 31 to 49 | 33 (44.6) |  | 0.77 (0.42, 1.42) 0.41 | 0.95 (0.47, 1.89) 0.88 |
| **Religion** |  |  |  |  |
| Christians | 89 (51.7) | 1.89 (0.17) | Ref | Ref |
| Muslims | 11 (37.9) |  | 1.755 (0.78, 3.94) 0.17 | 2.1 (0.86, 5.26) 0.10 |
| **Education** |  |  |  |  |
| None | 10 (55.6) | **6.56 (0.04)** | Ref | Ref |
| Primary | 49 (47.6) |  | 0.48 (0.16, 1.48) 0.20 | 0.39 (0.11, 1.30) 0.12 |
| SHS | 26 (65) |  | 0.66 (0.31, 1.40) 0.28 | 0.72 (0.33, 1.60) 0.42 |
| Tertiary | 15 (37.5) |  | 0.32 (0.13, 0.80) **0.015** | **0.39 (0.13, 0.99) 0.049** |
| **Residential** |  |  |  |  |
| Rural | 40 (67.8) | **10.97 (0.04)** | Ref | Ref |
| Peri-urban | 22 (44) |  | 0.37 (0.17, 0.82) **0.013** | 0.13 (0.02, 0.7) **0.02** |
| Urban | 38 (41.3) |  | 0.33 (0.17, 0.66) **0.02** | 0.10 (0.02, 0.78) **0.03** |
| **Trimester** |  |  |  |  |
| First | 13 (52) | 0.07 (0.97) | Ref | Ref |
| Second | 34 (50) |  | 1.12 (0.47, 2.69) 0.78 | 1.3 (0.51, 3.30) 0.58 |
| Third | 53 (49.1) |  | 1.04 (0.57, 1.90) 0.91 | 1.1 (0.58, 2.12) 0.75 |
| **Gravidity** |  |  |  |  |
| Gravida I | 25 (53.2) | 1.14 (0.57) | Ref | Ref |
| Gravida II | 25 (43.9) |  | 1.46 (0.67, 3.16) 0.344 | 1.37 (0.54, 3.48) 0.50 |
| >Gravida II | 50 (51.5) |  | 1.07 (0.53, 2.14) 0.85 | 0.99 (0.38, 2.62) 0.99 |

### Transmission Dynamics of *T. gondii* infection among Pregnant Women

**Table 4** shows that participants with backyard animals, with the extensive and semi-intensive system, without veterinary care, and contact with animal faeces had significant odds of being infected with *T. gondii* [AOR=3.90 (1.60 – 9.48) p=0.003], [AOR=31.7 (20.90 –8921.3) p< 0.001]; AOR=5.2 (3.37 – 367.70) p=0.003], [AOR=2.90 (1.10 – 7.46) p=0.031], and [AOR=3.90 (1.81 –8.35) p=0.001], respectively, whereas the use of disposable gloves to hand animals’ faeces significantly reduced the odds [AOR=0.05 (0.002– 0.91) p=0.043]. Table 4 further shows that participants using river, stream, or well water had 2.91 times the odds of being infected with *T. gondii* infections [AOR=2.91 (1.07 – 7.92) p=0.037], whereas participants who treated their water before drinking had less risk of the infection [AOR=0.26 (0.10 – 0.70) p=0.008]. There was no association between eating undercooked animal meat and drinking unpasteurized milk with *T. gondii* infections (**P-value > 0.05).**

**Table 4.** Associated Risk Factors for the transmission of *T*. *gondii* Infection among the study population.

| Risk factors | *T. gondii* Positive (%) | $\chi^2$ (P-value) | COR (95%CI) p-v | AOR (95%CI) p-v |
| --- | --- | --- | --- | --- |
| **Animal in household** |  |  |  |  |
| Yes | 92 (58.6) | 22.5 (0.001) | 6.4 (2.78, 14.60) **0.001** | **3.9** (1.60, 9.48) **0.003** |
| No | 8 (18.2) |  | Ref | Ref |
| **Type of animals** |  |  |  |  |
| Pet (cat, dog, etc.) | 51(56.7) | 0.5 (0.779) | 0.75 (0.29, 1.96) 0.55 | 0.96 (0.33, 2.81) 0.93 |
| Poultry/birds | 27(61.4) |  | 0.91 (0.32, 2.62) 0.86 | 1.13 (0.35, 3.66) 0.84 |
| Ruminant | 14(63.6) |  | Ref | Ref |
| **Level of confinement** |  |  |  |  |
| Extensive system | 71(94.7) | 92.1 (0.001) | 16.9(42.6, 653.6)**0.001** | **31.7**(20.9, 8921.3) **0.001** |
| Semi-intensive | 17(56.7) |  | 4.3 (3.81, 39.65) **0.001** | **5.2** (3.37, 367.7) **0.003** |
| Intensive system | 5(9.6) |  | Ref | Ref |
| **Contact with animal droppings** |  |  |  |  |
| Yes | 54(66.7) | 11.9 (**0.001**) | 3.4 (1.67, 6.78) **0.001** | **3.9** (1.81, 8.35) **0.001** |
| No | 22(37.3) |  | Ref | Ref |
| **Veterinary care** |  |  |  |  |
| Yes | 22(50) | 6.37 (**0.041**) | 1.3 (0.47, 3.59) 0.61 | 1.4 (0.45, 4.10) 0.51 |
| No | 60(67.4) |  | 2.7 (1.06, 6.86) **0.038** | **2.9** (1.10, 7.46) **0.031** |
| Do not know | 10(43.5) |  | Ref | Ref |
| **Use gloves to handle animal faeces.** |  |  |  |  |
| Yes | 2 (25) | 14.7 (**0.001**) | 0.07 (0.01, 0.38) **0.002** | **0.05** (0.002, 0.91) **0.043** |
| No | 90(82.6) |  | Ref | Ref |
| **Eating of animal meat** |  |  |  |  |
| Well cooked | 71(48.3) | 0.46 (0.497) | 0.81 (0.43, 1.51) 0.50 | 0.80 (0.43, 1.50) 0.49 |
| Undercooked or raw | 29 (53.7) |  | Ref |  |
| **Take unpasteurized milk** |  |  |  |  |
| No | 74(49.3) | 0.04 (0.839) | 0.94 (0.49, 1.77) 0.84 | 0.93 (0.50, 1.76) 0.83 |
| Yes | 26 (51) |  | Ref |  |
| **Primary source of drinking water** |  |  |  |  |
| Pipe | 45(39.5) | 14.9 (0.002) | 0.61 (0.28, 1.34) 0.22 | 0.60 (0.27, 1.34) 0.21 |
| Borehole | 7(58.3) |  | 1.3 (0.35, 5.01) 0.69 | 2.72 (0.60, 12.23) 0.19 |
| River/stream/well | 31(73.8) |  | 2.7 (1.01, 6.99) **0.049** | 2.91 (1.07, 7.92) **0.037** |
| Bottle/sachet water | 17(51.5) |  | Ref | Ref |
| **Treat water before drinking.** |  |  |  |  |
| Yes | 9(29) | 6.3 (0.012) | 0.36 (0.16, 0.82) **0.015** | 0.26 (0.10, 0.70) **0.008** |
| No | 91(53.5) |  | Ref | Ref |
| **Share water sources with animals.** |  |  |  |  |
| Yes | 19(76) | 7.9 (0.005) | 3.7(1.42, 9.74) **0.008** | 2.2 (0.76, 6.40) 0.14 |
| No | 81(46) |  | Ref |  |

(Data Source: Field Data and Laboratory records, 2023)
Note: P-v = P-Value; Ref = reference; CI = confidence interval.

### Association of Toxoplasmosis on Pregnancy Outcome

**Table 5** shows that pregnant women with *T. gondii* infection were associated with miscarriage (χ^2^=6.1, p= 0.014). Participants who had ever had a miscarriage were 2.93 times more likely to be infected with *T. gondii* [AOR=2.93 (1.20 – 7.04), p=0.016].

**Table 5.** Association of Toxoplasmosis on Pregnancy Outcomes.

| Risk factors | *T. gondii*<br>Positive (%) | $\chi^2$<br>(P-value) | COR (95%CI) p-v | AOR (95%CI) p-v |
| --- | --- | --- | --- | --- |
| **Child with hearing loss** |  |  |  |  |
| Yes | 1 (50) | 0.00 (1.0) | 1.0 (0.06, 16.3) 1.0 | 1.7 (0.10, 29.3) 0.72 |
| No | 70 (50) |  | Ref | Ref |
| **Miscarriage history** |  |  |  |  |
| miscarriage in past | 24 (66.7) | **6.1 (0.014)** | 2.6 (1.20, 5.75) **0.016** | 2.93 (1.2, 7.04) **0.016** |
| No miscarriage | 51 (43.2) |  | Ref | Ref |
| **Had chorioretinitis** |  |  |  |  |
| Yes | 34 (55.7) | 1.26 (0.263) | 1.4 (0.77, 2.58) 0.26 | 1.8 (0.82, 3.93) 0.14 |
| No | 66 (47.1) |  | Ref | Ref |

(Data Source: Field Data and Laboratory records, 2023)
*Note: Each response did not sum up to 201.*

## 4. Discussion

### Seroprevalence of *T. gondii*

The seroprevalence of *T. gondii* infection was 49.75% among pregnant women and aligns well with previous studies in Ghana, reporting between 50% and 56% in health facilities [20, 30, 31]. Meanwhile, other studies in Ghana have reported over 76% seroprevalence of *T. gondii* infection among pregnant women in health facilities, much higher than our study [32–34]. The seroprevalence rate in this study is much higher than other studies in Ghana and Ethiopia, which reported 21.5% [35] and 35.6% [36], respectively. However, similar studies in Western Romania and Brazil reported 55.8% and 71% [37, 38].

The disparities in the prevalence rates of *T. gondii* infections reported in these studies could be linked to the different settings, cultural practices, hygiene practices, environment, dietary habits, socio-economic conditions, and methodological approaches [39, 40]. This current study observed that participants residing in rural environs with low socio-economic status were vulnerable to *T. gondii* infection. This suggests that rural environments increase the risk of disease transmission.

This study further revealed that *T. gondii* infections were significantly associated with poor management of pets, backyard farm animals, veterinary services, and rural settlements [41, 42]. In this study, nearly 10% of the study participants had acute infection with *T. gondii*, of which about 3% had naïve immunity with the potential risks of mother-to-child transmission of the parasites [20, 43] with its dire consequences [20, 44].

### Prevalence of Congenital Infections (HBV, Syphilis and HIV) Screened at ANC among the Participants

Ghana, over the years, has formulated policies for screening expectant mothers for viral hepatitis, HIV/AIDS, syphilis, and malaria to improve pregnancy outcomes and neonatal health [15, 16]. Nevertheless, this study reported a high prevalence of 14.61%. 3.07% and 0.61% for hepatitis B, syphilis, and HIV infections, respectively, among the participants.

The rate of viral hepatitis B (HBV) infection was much higher than in previous studies in Ghana which reported between 2.4% and 10.6% among pregnant women [45–53]. These disparities in HBV prevalence among pregnant women in Ghana could be attributed to the different settings and cultural and behavioural characteristics of the participants [54]. In 2002, Ghana initiated a national expanded program for hepatitis B immunization [55] in response to the WHO target to eliminate HBV infection by 2030 [56, 57]. However, this current rate of HBV infection suggests a low vaccine uptake, and the 2030 target may not be achieved. This current rate presents a high risk for mother-to-child transmission, which is the main route of HBV infection, including sexual and other behavioural characteristics [55, 58].

The seroprevalence of syphilis was 3.07% among the participants, well aligned with the 3.7% and 4.1% reported among pregnant women at the Cape Coast Metropolitan Hospital [59] and gold miners in Konongo [60] in Ghana. This rate is, however, higher than 0.4%, previously reported in the general population [61]. Several studies have also reported syphilis among pregnant women: 2.5% in Tanzania [62], 1.9% in Ethiopia [63] and 4.4% in Brazil [64]. These prevalence rates somewhat differ from value reported in our study. The variations in these rates may be associated with several factors, including sexual behaviours, lifestyle factors, availability and access to screening services, and cultural and geographical settings [65].

This current study reported a 0.61% HIV infection rate, much lower than the 1.89%, 1.91%, and 1.66% among the adult population, in the Asante Mampong municipal, Ashanti region and the nation, respectively [66]. The current study result is at variance with 2.8% reported among pregnant women in the Asante Municipality in an HIV Sentinel survey [61]. The low HIV prevalence in this current study could be due to several causes, including the small number of participants screened, the sample size, and the data collection duration. This study observed a very low screening rate for HIV among the participants, and this can be a drawback for the zero-prevalence expected among mothers.

### Toxoplasmosis Concurrence with Congenital Infections Screened at ANC

This study reported 15%, 1%, and 4% *T. gondii* concurrence with HBV, HIV, and syphilis, respectively, among the population. However, this study did not establish an association between *T. gondii* co-infections with HBV, HIV, and syphilis, as reported earlier [67]. This study finding suggests that the co-occurrence infection of these diseases may be coincidental and thus did not influence the risk of *T. gondii* transmission. Hence, there is a need for independent screening and preventive strategies for these conditions in pregnancy. This finding in the current study, however, disagreed with an earlier study in Tanzania that reported that toxoplasmosis increases death rates among people living with HIV [68].

### Association of *T. gondii* infection with socio-demographic characteristics of participants

Although the prevalence of *T. gondii* decreases with increasing age in consonance, as in previous studies in Italy [69] and Pakistan [70], there was no association. However, this finding contradicts earlier reports in Nigeria [71], the USA [72], and the UK [73], which associated increasing age with *T. gondii* infection. The sharp difference could be attributable to our study’s generally younger population of women in their reproductive age.

The findings in this study demonstrated that respondents’ educational status and residential area were associated with *T. gondii* infection, as earlier reported [74]. Participants with primary or informal education had an increased risk of *T. gondii* infection in consonance with a report from Ethiopia [75]. Rural dwellers had an increased risk of infection than urban residents, probably due to the inadequate and deplorable social amenities and environmental factors [76]. The plausible reasons for the increased risk of *T. gondii* infection among participants with low education and rural residents may be due to their generally low socio-economic status, limited knowledge of the disease transmission, exposure to backyard animals and pets, and limited access to health facilities, and screening and treatment [77–79].

### Transmission Dynamics of *T. gondii* infection among Pregnant Women

This study has shown that the presence of backyard farm animals in households, their levels of confinement, exposure to faecal droppings, and veterinary care services were significant risk factors for *T. gondii* transmission. Cat is the primary host of *T. gondii,* with several other domestic animals as secondary and mechanical hosts [80], and their interaction with humans can transmit the parasites through their fur, saliva, or faeces [81]. This assertion is affirmed by previous studies that implicated the interaction of domesticated cats and dogs as a risk of *T. gondii* transmission to humans [82, 83]. This study also revealed that backyard animal husbandry practices increase the risk of *T. gondii* infection. The free-range backyard animal-rearing practice is more associated with rural households with little or no veterinary care, increasing the risk of *T*. *gondii* transmission. These animals could contaminate the environment with their faeces, which contain parasites and pose a risk to humans. In this study, contact with animal faeces increased the risk of *T. gondii* infection four-fold. This implies that droppings and litter from animals in the environment and poor hygiene practices can facilitate the transmission of *T. gondii* to humans [84]. However, using personal protective equipment to handle animals’ faeces highly reduced the risks of *T. gondi*i infection.

In this study, the consumption of animal products was not associated with *T. gondi*i infection, contrary to an earlier report [67] that linked eating meat and other animal products to the infection [85]. This study has shown that there is a three-fold increased risk of *T. gondii* infection in drinking water from rivers, streams, or wells, akin to a study in Brazil [86]. Livestock often share rivers and streams with humans and are most likely contaminated with faecal droppings containing *T. gondii* oocysts. The tachyzoite and bradyzoite forms of the parasite can persist in water and be accidentally ingested. However, treating water before drinking decreases the risk of *T. gondii* infection.

### Effect of Toxoplasmosis on Pregnancy Outcomes

This study has indicated that *T. gondii* infection was significantly associated with a higher risk of miscarriage among participants. Participants who were seropositive for toxoplasmosis had a threefold increased risk of miscarriages. *T. gondii* infection increases the risk of miscarriage due to its invasion of the placenta, causing inflammation and damage and leading to spontaneous expulsion of the fetuses [12]. This assertion aligns with several studies that linked *T. gondii* infection with spontaneous abortion [87–89]. The study findings highlight the public health significance and call for concerted efforts to integrate *T. gondii* screening with other congenital diseases to improve pregnancy outcomes.

### Limitation of the Study

The sample size of this study was relatively small compared to the study population and may fail to capture the diversity of the population. The serological method was only used to determine the status of *T. gondii* infection without confirmatory tests, such as PCR or avidity tests, with its inherent limitations. The study participants’ responses may suffer from recall bias, compromising their reports. This was a health facility-based study and could influence participants’ responses. The test results of the congenital diseases were based on those reported by the facility and were subject to inaccuracies. Notwithstanding these limitations, the outcomes of this study are relevant for public health policy consideration that could improve maternal and neonatal healthcare services.

## 5. Conclusion

The burden of toxoplasmosis and concurrence with HBV and syphilis was high among pregnant women, but they were not at risk for its transmission. Low levels of education, living in rural areas, animals in households, little or no veterinary services, and drinking water from rivers significantly influenced the transmission of *T. gondii*. An infection with *T. gondii* in pregnancy significantly increases the risk of miscarriage. The high prevalence and risks of *T. gondii* infections call for an urgent need to integrate toxoplasmosis into the ANC routine screening to improve pregnancy outcomes.

## Data Availability

All data produced in the present study are available upon reasonable request to the authors

## Acknowledgements

We sincerely acknowledge the invaluable contributions of Olivia Ndele, Uginabor Jonathan Matani, Lambon Isaac, Tikpinja Moses, Bright Bindink Kididisil, Beatrice Ellen, and Adam Kofi Jamiru, former Public Health students of the Akenten Appiah-Menka University of Skills Training and Entrepreneurial Development (AAMUSTED), Asante Mampong Campus. Their dedication to data collection and entry of responses into Google Forms during their voluntary attachment at the health facilities was instrumental to this study. We are also deeply grateful to Mr. Richard Sombeley Assoah and Madam Ndaayaa Margaret Kweasi, the brother and mother of the lead author (Ebenezer Assoah), respectively, for their support throughout the course of this study.

## Availability of supporting data

All data used for this manuscript are available upon a reasonable request.

## Author(s) information Affiliations

1Department of Biological Sciences Education, Akenten Appiah-Menka University of Skills Training and Entrepreneurial Development (AAMUSTED), Asante Mampong Campus, Ghana

2Department of Public Health Education, Akenten Appiah-Menka University of Skills Training and Entrepreneurial Development (AAMUSTED), Asante Mampong Campus, Ghana

3Department of Theoretical and Applied Biology, Kwame Nkrumah University of Science and Technology, Kumasi, Ghana

4Department of Science Education, Seventh–Day Adventist College of Education, Agona-Ashanti, Ghana

5Department of Science and Information Communication Technology, Effiduase Senior High Technical School, Ashanti Region, Ghana

## The Corresponding author*

^1^Department of Biological Sciences Education, Akenten Appiah-Menka University of Skills Training and Entrepreneurial Development (AAMUSTED), Asante Mampong Campus, Ghana.

## Authors Contributions

EA was responsible for data collection, data analysis, and drafting of the manuscript. DDY played a pivotal role in conceptualizing and developing the manuscript, provided critical edits, contributed to data analysis and interpretation, and ensured the manuscript’s intellectual rigor and clarity. PKA reviewed the manuscript and approved it for publication. GIB reviewed and structured the manuscript for publication. RA participated in data collection and reviewed the manuscript. JKZ managed the data, reviewed the manuscript, and approved it for publication. All authors have thoroughly read and approved the final manuscript for publication. EA and DDY are the joint senior authors for this manuscript with equal contributions, from conceptualization to the final approval of the work for publication.

## Ethics declarations

Ethical clearance was acquired from the Committee on Human Research, Publications, and Ethics (CHRPE) at the Kwame Nkrumah University of Science and Technology (with reference number CHRPE/AP/717/23).

## Ethics approval and consent to participate

Written permissions were obtained from the Ashanti Regional and Asante Mampong Municipal Health Directorates and the study facilities. Before sample collection, each study participant signed an informed consent form.

## Consent for publication

We (Authors) have all read the final manuscript and consented to it for publication.

